# Perioperative transcutaneous auricular vagus nerve stimulation for the prevention of postoperative nausea and vomiting after gynecologic laparoscopic surgery: protocol for a randomized, double-blind, sham-controlled trial

**DOI:** 10.64898/2026.09.24.26363854

**Authors:** Ru Wang, Hua Yang, Hui-Juan Li, Yi-Ran Yang, Jia-Le Du, Yan-Wen Zhou, Jian-Jun Yang, Long He

**Affiliations:** Department of Anesthesiology, Pain and Perioperative Medicine, The First Affiliated Hospital of Zhengzhou University, Zhengzhou 450000, Henan, China

**Keywords:** postoperative nausea and vomiting, transcutaneous auricular vagus nerve stimulation, gynecologic laparoscopic surgery, perioperative medicine, randomized controlled trial

## Abstract

**Background:** Postoperative nausea and vomiting (PONV) remains a common and distressing complication after gynecologic laparoscopic surgery, despite the use of multimodal antiemetic prophylaxis. Transcutaneous auricular vagus nerve stimulation (taVNS) is a noninvasive neuromodulation technique that targets vagal afferents in the external ear and may modulate brainstem and autonomic pathways involved in nausea and vomiting. This randomized trial will evaluate taVNS as an adjunctive, nonpharmacological strategy for preventing PONV in women undergoing gynecologic laparoscopic surgery.

**Methods:** This single-center, prospective, randomized, double-blind, sham-controlled, parallel-group superiority trial will enroll 146 adult women undergoing elective gynecologic laparoscopic surgery under general anesthesia. Participants will be randomly assigned in a 1:1 ratio to receive active taVNS or sham stimulation, with randomization stratified by Apfel risk score (≤2 vs ≥3). Participants will receive two 30-minute stimulation sessions: one beginning 30 minutes before induction of anesthesia and the other beginning 30 minutes after arrival in the post-anesthesia care unit. The primary outcome is the proportion of participants experiencing PONV during the first 48 hours after surgery, according to the prespecified definition. Secondary outcomes include nausea severity, time to the first PONV episode, the number of vomiting or retching episodes, rescue antiemetic use, postoperative pain intensity, cumulative opioid consumption expressed as morphine milligram equivalents, Quality of Recovery-15 scores at 24 and 48 hours, and prespecified measures of gastrointestinal recovery. The primary analysis will follow the intention-to-treat principle. A per-protocol analysis and prespecified missing-data sensitivity analyses will also be performed.

**Conclusions:** This trial will evaluate the efficacy and safety of perioperative taVNS for PONV prevention in women undergoing gynecologic laparoscopic surgery. If effective, taVNS may represent a feasible nonpharmacological adjunct to standard antiemetic prophylaxis and may be incorporated into enhanced recovery after surgery pathways.

**Trial registration:** Chinese Clinical Trial Registry, ChiCTR2600115965; registered on 4 January 2026.

## Introduction

Postoperative nausea and vomiting (PONV) is one of the most common and distressing complications following general anesthesia. It affects approximately 25%– 30% of surgical patients and may occur in as many as 70%–80% of patients at high-risk [1]. Although PONV is rarely life-threatening, it can substantially impair patient comfort, delay discharge from the post-anesthesia care unit (PACU), and increase the risk of unplanned hospital admissions[2].

Established risk factors for PONV include female sex, a history of motion sickness or previous PONV, nonsmoking status, and younger age[3]. Gynecologic and laparoscopic procedures are associated with an especially high risk because of pneumoperitoneum, visceral manipulation, and possible hormonal or autonomic influences. Exposure to volatile anesthetics, nitrous oxide, and perioperative opioids may further increase the emetogenic burden, making women undergoing gynecologic laparoscopic surgery a particularly vulnerable population [4].

Pharmacologic prophylaxis, most commonly involving combinations of 5-hydroxytryptamine-3 (5-HT₃) receptor antagonists, neurokinin-1 (NK₁) receptor antagonists, and corticosteroids, remains the cornerstone of PONV prevention[5]. Nevertheless, complete prevention is not achieved in a considerable proportion of patients, and the residual incidence of PONV remain approximately 30%–40% during the first 24 hours after surgery. Antiemetic drugs may also cause adverse effects, including headache, dizziness, sedation, and QT-interval prolongation [6]. These limitations highlight the need for effective and safe nonpharmacologic adjuncts.

Current guidelines recommend several complementary strategies for reducing PONV risk, including adequate hydration, carbohydrate loading, aromatherapy, and stimulation of the Pericardium-6 (PC6) acupuncture point [1]. PC6 stimulation has demonstrated antiemetic efficacy in various perioperative settings. However, conventional acupuncture and acupoint injection may be invasive and difficult to implement routinely [7]. Transcutaneous electrical acupoint stimulation offers a noninvasive alternative and has shown potential benefits in reducing PONV, although the underlying mechanisms and optimal stimulation parameters remain uncertain [8].

The vagus nerve plays an important role in the gut–brain axis by transmitting afferent signals from visceral organs to brainstem nuclei, including the nucleus tractus solitarius and the dorsal vagal complex, which participate in the integration of emetic reflexes [9]. The effects of several antiemetic interventions on vagally related brainstem pathways support the possibility that modulation of vagal activity may help reduce nausea and vomiting [10].

Transcutaneous auricular vagus nerve stimulation (taVNS) delivers mild electrical impulses to regions of the external ear innervated, at least in part, by the auricular branch of the vagus nerve (ABVN), including the cymba conchae and cavum conchae. Functional neuroimaging studies suggest that taVNS can modulate brainstem and cortical regions involved in autonomic regulation without the surgical risks associated with implanted vagus nerve stimulation [11]. taVNS exerts multifaceted effects that encompass both physiological regulation and clinical symptom modulation. It enhances parasympathetic activity and restores autonomic balance[12–14], activates the cholinergic anti-inflammatory pathway to suppress peripheral and central inflammatory responses[15, 16], In addition, taVNS regulates the brain–gut axis by influencing visceral afferent signaling and gastrointestinal motility[17, 18]. Collectively, these mechanisms contribute to clinical benefits such as alleviation of visceral discomfort and symptom improvement in disorders like irritable bowel syndrome, which may share pathophysiological mechanisms relevant to the prevention of PONV.

On this physiological basis, perioperative taVNS may be a safe and promising adjunct for reducing PONV by modulating vagal afferent activity and central autonomic function. We therefore designed this randomized, double-blind, sham-controlled trial to evaluate the efficacy and safety of taVNS administered before anesthesia induction and during early postoperative recovery in women undergoing gynecologic laparoscopic surgery.

## Methods

### Study design and setting

This study is a single center, prospective, randomized, double-blind, sham-controlled, parallel-group superiority trial conducted at the First Affiliated Hospital of Zhengzhou University. The protocol was developed in accordance with the Standard Protocol Items: Recommendations for Interventional Trials (SPIRIT) guidelines, and the final report will be prepared in accordance with the Consolidated Standards of Reporting Trials (CONSORT) statement.

The planned recruitment period is from December 1, 2025, to December 31, 2026.

The trial was registered in the Chinese Clinical Trial Registry under registration number ChiCTR2600115965.

### Eligibility Criteria Inclusion criteria

Female patients aged ≥18 years; scheduled for elective laparoscopic gynecologic surgery (e.g., ovarian cystectomy, salpingectomy, myomectomy, or hysterectomy) under general anesthesia; classified as American Society of Anesthesiologists (ASA) physical status I - Ⅲ.; able to understand the study procedures and provide written informed consent.

### Exclusion criteria

Patients will be excluded if they meet any of the following criteria: Allergy to or contraindication for any study medications; Chronic or long-term use of antiemetic, emetogenic, opioid, or glucocorticoid medications; A history of chronic pain, psychiatric disorders, epilepsy, or use of centrally acting medications that could interfere with neuromodulation or outcome assessment; Presence of a cardiac pacemakers, implantable cardioverter-defibrillators, cochlear implants, or other electronic or metallic implants that could be affected by electrical stimulation. Skin lesions, infection, inflammation, or deformity at or near the auricular stimulation sites; Significant cardiovascular disease, including clinically relevant arrhythmia, bradycardia, severe hypertension, or heart failure; Pregnancy or breastfeeding; Current participation in another interventional study. Inability to communicate, cooperate, or comply with study procedures.

### Recruitment, informed consent, and withdrawal

Potentially eligible patients will be screened during preoperative assessment. Written informed consent will be obtained by a trained investigator before any study-specific procedure is performed. Participants may withdraw from the study at any time without affecting their medical care. Prespecified reasons for withdrawal include major protocol violations, conversion from laparoscopic to open surgery, prolonged postoperative mechanical ventilation, admission to the intensive care unit, or any clinical circumstance that prevents completion of the intervention or follow-up assessments. Data collected before withdrawal will be included in the intention-to-treat analysis unless the participant explicitly requests that their data not be used, where permitted by applicable regulations and the ethics approval.

### Randomization and blinding

Participants will be randomly assigned in a 1:1 ratio to either the active taVNS group or the sham-stimulation group. The randomization sequence will be generated by an independent member of the research team using the Sealed Envelope randomization service (https://www.sealedenvelope.com/simple-randomiser/v1/lists). Permuted blocks of variable size, including blocks of 2 and 4, will be used. Randomization will be stratified by Apfel risk score (≤2 versus ≥3) to promote balance in baseline PONV risk.

Group assignments will be concealed using sequentially numbered, sealed, opaque envelopes, which will be opened immediately before the intervention by a research staff member not involved in patient care or outcome assessment. Blinding will be maintained at multiple levels. Participants, anesthesiologists, post-anesthesia care unit (PACU) personnel, ward nurses, data collectors, and statisticians will all remain blinded to group allocation. Only a dedicated interventionist, who is not involved in anesthesia management or outcome assessment, will perform the stimulation procedures.

Allocation concealment will be maintained using sequentially numbered, sealed, opaque envelopes. The envelope will be opened immediately before the intervention by a designated research staff member who is not involved in anesthesia management or outcome assessment. Participants, anesthesiologists, PACU personnel, ward nurses, outcome assessors, data collectors, and statisticians will remain blinded to treatment allocation. Only the interventionist responsible for delivering the stimulation will be involved in the treatment procedure. The taVNS device will use coded treatment programs for active and sham stimulation. The two programs will be configured before trial initiation and labeled only as “Program A” and “Program B” . The device interface will not display the treatment assignment. The interventionist will be trained in device operation and safety procedures and will undergo competency assessment before participating in the trial. Both groups will use identical hardware and electrode placement. The sham condition will use a subperceptual stimulation protocol or no effective electrical current, according to the validated device configuration.

## Study interventions

### Device and stimulation parameters

A portable, battery-powered transcutaneous electrical stimulation device will be used. Electrodes will be placed at the cymba conchae, an auricular region containing cutaneous innervation from the auricular branch of the vagus nerve. Both groups will receive stimulation using biphasic square-wave pulses at a frequency of 25 Hz and a pulse width of 250 μs. In the active taVNS group, the current intensity will be gradually increased to a comfortable, clearly perceptible tingling sensation without pain, excessive discomfort, or visible muscle twitching. The intensity will then be maintained throughout the 30-minute session. The expected current intensity will generally range from 1 to 3 mA, although the final intensity will be individually adjusted according to participant comfort.

In the active sham group, the electrodes will be placed at the same auricular sites and the same waveform, frequency, pulse width, device interface, and session duration will be used. The sham current will be delivered according to a prespecified ramp-down program: the intensity will increase linearly from 0 to 0.5 mA during the first 30 seconds, increase from 0.5 to 1.0 mA during the subsequent 30 seconds, and then decrease linearly from 1.0 mA to 0 mA over the following 30 seconds. The current will remain at 0 mA for the remainder of the 30-minute session, while the electrodes remain in place. This procedure is intended to reproduce the initial sensory experience of electrical stimulation while avoiding sustained stimulation at the therapeutic intensity used in the active taVNS group. The possibility that brief initial stimulation may have residual biological effects cannot be completely excluded.

### Timing of Intervention

Each participant will receive two 30-minute stimulation sessions: The first session will begin 30 minutes before induction of anesthesia; The second session will begin 30 minutes after arrival in the PACU.

### Anesthetic and perioperative management

All participants will fast for 6–8 hours before surgery, and no routine premedication will be administered. Upon arrival in the preoperative holding area, baseline blood pressure and heart rate will be recorded. Standard intraoperative monitoring will include electrocardiography, noninvasive blood pressure, pulse oximetry, and bispectral index (BIS). After preoxygenation with 100% oxygen, anesthesia will be induced with midazolam 0.04 mg·kg⁻¹ or remimazolam 0.2 mg·kg⁻¹, propofol 1.5 mg·kg⁻¹, sufentanil 0.3 µg·kg⁻¹, and rocuronium 0.6 mg·kg⁻¹. A laryngeal mask airway will then be inserted. After successful airway placement, dexamethasone 5 mg intravenously will be administered for PONV prophylaxis. Mechanical ventilation will be initiated using volume-controlled ventilation. Tidal volume will be set at 6–8 mL·kg⁻¹ of predicted body weight, and respiratory rate will be adjusted to maintain end-tidal carbon dioxide 35-45 mmHg. Positive end-expiratory pressure will be set 5-8 cmH₂O. The fraction of inspired oxygen will be adjusted to maintain SpO₂ ≥95%. Anesthesia will be maintained with propofol 2–6 mg·kg⁻¹·h⁻¹ and remifentanil 4–10 µg·kg⁻¹·h⁻¹, supplemented with sevoflurane at 0.8–1.0 minimum alveolar concentration. The BIS will be maintained between 40 and 60. Mean arterial pressure will be maintained within ±20% of the baseline values. Ephedrine will be administered as clinically indicated for hypotension, and additional neuromuscular blocking agent will be administered when necessary to maintain adequate surgical conditions.

Approximately 10-15 minutes before the end of surgery, metoclopramide 10 mg intravenously will be administered as an additional antiemetic. At the end of surgery, anesthetic administration will be discontinued. After recovery of spontaneous respiration and airway reflexes, the LMA will be removed. Sugammadex 2-4 mg·kg⁻¹ will be administered when clinically indicated to reverse residual neuromuscular blockade. Patients will then be transferred to the post-anesthesia care unit (PACU) for standardized postoperative monitoring. A modified Aldrete score of at least 9 will be required for PACU discharge to the surgical ward. Palonosetron 0.25 mg intravenously will be administered as a rescue antiemetic therapy if nausea or vomiting occurs despite prophylaxis.

All patients will receive flurbiprofen axetil 50 mg intravenously intraoperatively. At the conclusion of surgery, before skin closure local anesthetic infiltration will be performed using 3-5 mL of 0.5% ropivacaine per incision site. Intravenous patient-controlled intravenous analgesia (PCIA) will consist of sufentanil 100 µg diluted with normal saline to a total volume of 100 mL, corresponding to a concentration of 1 µg/mL. The PCIA device will deliver a background infusion of 2 mL·h⁻¹, a bolus dose of 0.5 mL, and a lockout interval of 15 minutes. Postoperative pain will be evaluated using an 11-point numerical rating scale, ranging from 0 (no pain) to 10 (worst imaginable pain). If the NRS score remains ≥4 despite PCIA, rescue analgesia with oxycodone 2–5 mg intravenously will be administered.

### Schedule of enrollment, interventions, and assessments

Eligible participants will undergo screening and baseline assessment before randomization. After randomization, participants will receive either active taVNS or sham stimulation according to their assigned group. The preoperative stimulation session will begin 30 minutes before anesthesia induction, and the postoperative stimulation session will begin 30 minutes after arrival in the post-anesthesia care unit. Postoperative assessment times will be measured from the end of surgery, defined as completion of the final surgical skin closure. Each stimulation session will last 30 minutes. The schedule of enrollment, interventions, and assessments is summarized in Figure 1.

**Figure 1.**
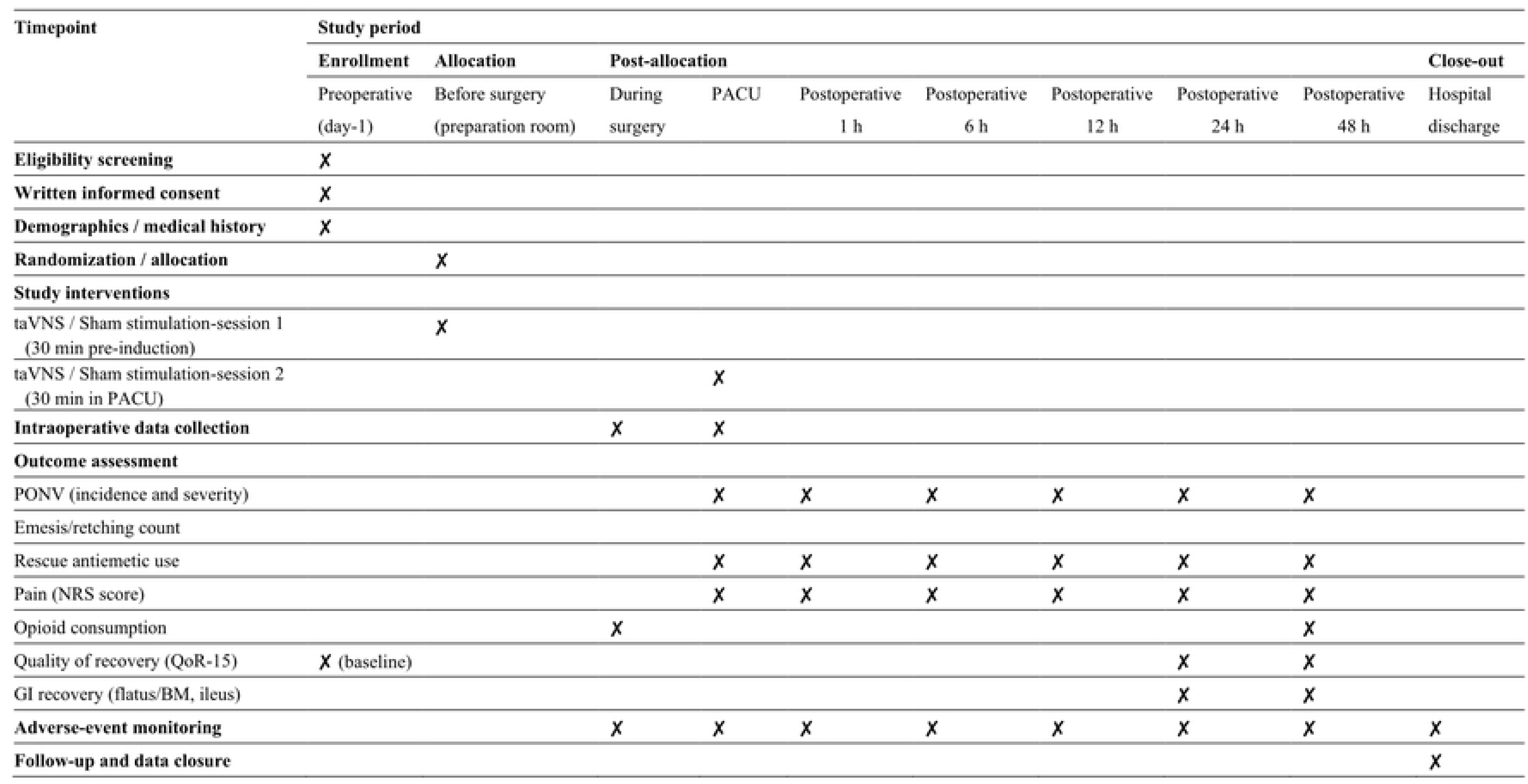
Schedule of enrollment, interventions, and assessments. The schedule follows the SPIRIT framework. Postoperative assessment times are measured from the end of surgery, defined as completion of the final surgical skin closure. Each stimulation session lasts 30 minutes. The preoperative stimulation session begins 30 minutes before anesthesia induction, and the postoperative stimulation session begins 30 minutes after arrival in the post-anesthesia care unit (PACU). The actual interval between the end of surgery and initiation of the postoperative stimulation session will be recorded.

### Participant flow and retention

Potentially eligible participants will be identified during the screening process and assessed against the predefined eligibility criteria. Eligible participants who provide informed consent will be randomized in a 1:1 ratio to receive active taVNS or sham stimulation. Participants will then undergo the allocated intervention and scheduled follow-up assessments. Reasons for exclusion, intervention discontinuation, and loss to follow-up will be recorded throughout the trial. The planned participant flow from screening through randomization, intervention, follow-up, and analysis is shown in Figure 2.

**Figure 2.**
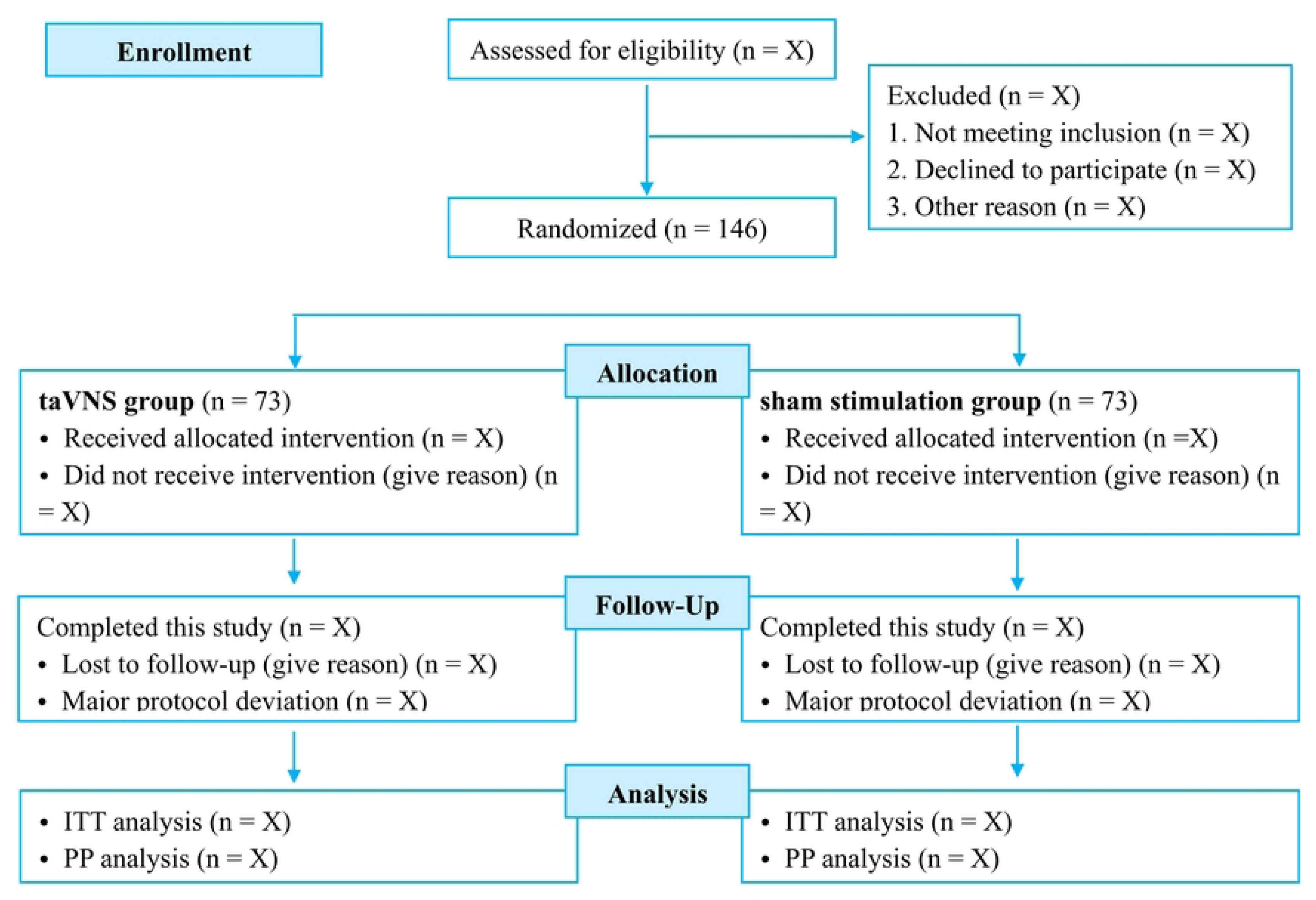
Planned participant flow through screening, randomization, intervention, follow-up, and analysis. A total of 146 participants are planned to be randomized in a 1:1 ratio to receive active transcutaneous auricular vagus nerve stimulation (taVNS) or sham stimulation. The actual numbers of participants and the reasons for exclusion, discontinuation of the intervention, and loss to follow-up will be reported in the completed trial report.

Strategies to promote retention and complete follow-up. To maximize retention, participants will receive standardized discharge instructions that include a 24-hour contact number for the research team, and participants discharged before completion of the 48-hour assessments will be contacted by telephone at the scheduled assessment times using standardized questionnaires. For participants who discontinue the intervention or withdraw consent, all data collected up to that point will be retained and analyzed as prespecified; with the participant’s consent, the primary outcome (PONV within 48 hours), safety data, nausea severity and pain scores, QoR-15 scores, and opioid consumption will continue to be collected by telephone where feasible. A list of the outcome data to be collected from participants who discontinue is therefore: PONV status and episodes, nausea severity, pain intensity, rescue antiemetic and rescue analgesic use, opioid consumption, QoR-15 score, gastrointestinal recovery, and adverse events

### Outcomes Primary Outcome

The primary outcome is the proportion of participants who experience postoperative nausea and vomiting (PONV) within 48 hours after surgery. For this outcome, PONV is defined as the occurrence of any nausea, retching, or vomiting during the observation period. Nausea alone will therefore qualify as a primary outcome event.

Nausea is defined as an unpleasant subjective sensation associated with the urge to vomit. Retching is defined as an involuntary attempt to vomit without the expulsion of gastric contents, and vomiting is defined as the forceful expulsion of gastric contents through the mouth. A trained investigator blinded to treatment allocation will assess PONV at 1, 6, 12, 24, and 48 hours after surgery. At each assessment, participants will be asked about symptoms occurring since the previous assessment, rather than only symptoms present at the assessment time. Participant reports will be supplemented by available nursing records and records of rescue antiemetic administration. Each participant will be classified as having or not having experienced PONV during the entire 48-hour observation period, irrespective of the number of episodes or the use of rescue antiemetics.

### Secondary Outcomes

1 Nausea severity. Nausea severity will be assessed using an 11-point numerical rating scale (NRS), ranging from 0 (no nausea) to 10 (worst imaginable nausea), at 1, 6, 12, 24, and 48 hours after surgery.

2 Time to first PONV episode. The elapsed time from the prespecified postoperative time origin to the first reported episode of nausea, retching, or vomiting will be recorded.

3 Vomiting and retching episodes. The cumulative number of vomiting and retching episodes within 48 hours will be recorded.

4 Rescue antiemetic use. Outcomes will include the proportion of participants receiving rescue antiemetics and the type, number of administrations, and cumulative dose of each rescue antiemetic within 48 hours. Doses of different antiemetic drugs will be reported separately.

5 Postoperative pain. Pain intensity will be assessed using an 11-point NRS, ranging from 0 (no pain) to 10 (worst imaginable pain), at 1, 6, 12, 24, and 48 hours after surgery.

6 Postoperative opioid consumption. Total opioid consumption within 48 hours will include opioids delivered through patient-controlled intravenous analgesia and any additional rescue opioids. Consumption will be reported for each opioid and converted to a common morphine-equivalent metric using prespecified, route-specific conversion factors.

7 Quality of recovery. Quality of recovery will be assessed using the Quality of Recovery-15 (QoR-15) questionnaire on the day before surgery and at 24 and 48 hours after surgery. Total scores range from 0 to 150, with higher scores indicating better recovery. A validated language version will be used.

8 Gastrointestinal recovery. Time to first flatus and time to first bowel movement will be recorded separately. The original protocol also includes postoperative ileus within 48 hours; its diagnostic criteria and ascertainment procedure require perspectivization.

### Data Collection and Management

Study data will be collected using standardized case report forms (CRFs) during the preoperative, intraoperative, and postoperative periods. Investigators responsible for outcome assessment will receive standardized training and will remain blinded to treatment allocation. Preoperative data will include demographic characteristics, relevant medical history, American Society of Anesthesiologists physical status, planned surgical procedure, the individual components of the simplified Apfel risk score, and the baseline QoR-15 score. The simplified Apfel score comprises female sex, nonsmoking status, a history of PONV or motion sickness, and anticipated postoperative opioid use. Intraoperative data will include the surgical procedure performed, anesthesia and surgery durations, airway management, anesthetic and analgesic medications and doses, prophylactic antiemetic administration, and clinically relevant physiological measurements and adverse events. Intervention records will document the timing and duration of each stimulation session, completion status, interruptions, and reasons for incomplete treatment. Device settings and delivered stimulation parameters will be retained in a restricted-access record where disclosure could compromise blinding. Postoperative outcomes will be collected at 1, 6, 12, 24, and 48 hours after surgery. Data will be obtained from participant interviews and relevant clinical records. Participants discharged before completion of the 48-hour observation period will be contacted by telephone using standardized questions. The assessment method and timing will be documented. Each participant will be assigned a unique study identification number. Identifiable information will be stored separately from the research dataset. Paper records will be kept in locked cabinets, and electronic records will be stored on encrypted, password-protected institutional systems. Access will be restricted to authorized personnel according to their study responsibilities. Data quality will be maintained through checks for completeness, valid ranges, and internal consistency. Discrepancies will be resolved against source records, and corrections will be documented in an audit trail. Protocol deviations and reasons for missing data will be recorded. The principal investigator will oversee data quality and protocol adherence. The analysis dataset will be finalized after data queries have been resolved. Treatment codes will remain masked to the statistical analyst until the prespecified analysis procedures and data-cleaning decisions have been finalized.

### Sample Size Calculation

Based on preliminary institutional data from patients undergoing similar procedures, the 48-h incidence of PONV despite standard prophylaxis was estimated to be approximately 42%. We hypothesized that perioperative taVNS would reduce this incidence to 20%, representing a 22% absolute reduction. Based on a two-sided chi-square test with α = 0.05 and power = 0.80, 66 participants per group were required. Allowing a 10% dropout rate, the final sample size was set at 146 participants (73 per group). Sample size calculations were performed using PASS version 21.0 (NCSS, LLC, Kaysville).

### Statistical Analysis

Continuous variables will be summarized as means and standard deviations or medians and interquartile ranges, as appropriate. Categorical variables will be summarized as counts and percentages. Baseline characteristics will be described by randomized group without significance testing.

### Primary outcome analysis

The incidence of PONV within 48 hours will be summarized for each group. The primary treatment comparison will use a modified Poisson regression model with robust standard errors, including treatment group and the Apfel risk stratum used for randomization as covariates. The treatment effect will be reported as an adjusted risk ratio with a 95% confidence interval. An unadjusted risk ratio and absolute risk difference, with corresponding 95% confidence intervals, will also be reported. A two-sided P value below 0.05 will be considered statistically significant for the primary outcome.

### Secondary outcome analyses

Binary secondary outcomes, including receipt of rescue antiemetics, will be summarized by group and analyzed using regression methods appropriate to the event frequency. The number of vomiting or retching episodes will be analyzed using Poisson regression. Negative binomial regression will be used if substantial overdispersion is present. The selected model and any limitations arising from sparse data will be reported. Repeated nausea and pain scores will be analyzed using mixed-effects models incorporating treatment group, assessment time, and the group-by-time interaction. Time will be treated as a categorical variable, and within-participant correlation will be accounted for. Model assumptions will be assessed; an ordinal mixed-effects approach will be considered if a linear model is unsuitable for the observed score distribution. Postoperative QoR-15 scores will be analyzed using a repeated-measures model adjusted for the baseline QoR-15 score. Estimated between-group differences at 24 and 48 hours will be reported with 95% confidence intervals. Total postoperative opioid consumption will be compared using an appropriate generalized linear model or a nonparametric method, depending on its distribution.

Time to first PONV, first flatus, and first bowel movement will be analyzed as time-to-event outcomes. Participants without an observed event will be censored at the end of follow-up or their last reliable event-free assessment. Kaplan–Meier estimates and, where appropriate, Cox regression will be used if sufficiently precise event times are available. If event times are known only within assessment intervals, interval-censored methods will be considered.

### Missing data and sensitivity analyses

The amount, pattern, and reasons for missing data will be described by group. Missing primary outcome data will be addressed using multiple imputation under a missing-at-random assumption, where appropriate. The imputation model will include treatment allocation, randomization stratum, relevant baseline predictors, and available postoperative outcome information. For the composite primary outcome, any documented episode of nausea, retching, or vomiting within 48 hours will establish a positive outcome even if later assessments are missing. Participants with incomplete follow-up and no documented event will not automatically be classified as free of PONV. Sensitivity analyses will include complete-case and per-protocol analyses and analyses exploring departures from the missing-at-random assumption.

### Safety monitoring and adverse events

taVNS is generally well tolerated; reported adverse effects are typically mild and transient (e.g., ear discomfort/tingling). We will monitor bradycardia, hypotension, syncope, skin irritation/burn, and other unexpected events during stimulation and up to 48 h. Serious adverse events (SAEs) are reported to the ethics committee per institutional policy; stimulation is paused/terminated if clinically indicated.

### Ethical considerations

The study complies with the Declaration of Helsinki and ICH-GCP. Approval from the Ethics Committee of the First Affiliated Hospital of Zhengzhou University (Approval No. 2025-KY-1347-001) has been obtained. Written informed consent will be obtained from all participants. Findings will be disseminated via peer-reviewed journals and scientific meetings; de-identified data may be shared upon reasonable request following publication and in line with institutional approvals.

### Patient and public involvement

Patients and members of the public were not involved in the design of this study. A plain-language summary of the findings will be made available to participants upon request after publication.

### Trial status

Protocol version: 1.0, Aug-03-2025

Recruitment status at manuscript submission: Recruiting Date of first participant enrollment: Jun-05-2026 Anticipated recruitment completion date: Dec-31-2026

## Discussion

PONV remains an important barrier to recovery after gynecologic laparoscopic surgery. This population has several established risk factors, including female sex and exposure to postoperative opioids, while volatile anesthetic exposure and the surgical context may further contribute to risk. Despite prophylactic antiemetic treatment, some patients continue to experience clinically important symptoms. Additional nonpharmacological interventions may therefore be useful as components of multimodal prevention strategies [19]. To date, there is no clinical evidence evaluating the effect of taVNS, a novel non-invasive neuromodulation technique, on the prevention of PONV in this surgical population. The present study aims to evaluate the efficacy of perioperative taVNS in reducing the incidence and severity of PONV, as well as its effects on postoperative pain, opioid consumption, recovery quality, and gastrointestinal function in patients undergoing gynecologic laparoscopic surgery.

taVNS provides a safe and reversible method to activate afferent fibers of the ABVN, located primarily on the inner surface of the tragus and cymba concha. fMRI studies have demonstrated that taVNS modulates activity in brainstem nuclei involved in vagal regulation, such as the nucleus tractus solitarius and dorsal motor nucleus, which are closely linked to nausea and vomiting pathways[20, 21]. Experimental and clinical evidence suggests that taVNS enhances parasympathetic tone, suppresses central emetic signaling through modulation of serotonin and substance P release, and exerts anti-inflammatory effects[22, 23]. Preclinical models and clinical studies in irritable bowel syndrome have shown that taVNS improves gastrointestinal function and attenuates visceral pain[18, 24–26]. supporting its potential role as an adjunct in perioperative care.

The dual-session design of the present study—administering taVNS before anesthesia induction and again in the post-anesthesia care unit—was selected to maximize neuromodulatory coverage during the two periods. Preoperative stimulation may precondition vagal circuits and stabilize autonomic balance, while postoperative stimulation may reinforce vagal modulation during anesthetic washout, pain onset, and early mobilization. Our sham-controlled, double-blind approach aims to maintain methodological rigor while minimizing bias. To preserve blinding integrity, sham stimulation will be delivered at the same auricular site with sub-threshold current intensity, as relocating electrodes to non-vagal areas or using low-frequency stimulation could inadvertently activate vagal fibers or compromise blinding.

Beyond its potential antiemetic effect, taVNS may confer additional perioperative benefits through multiple physiological pathways. By enhancing parasympathetic activity and reducing systemic inflammation, taVNS could alleviate postoperative pain and decrease opioid requirements, consistent with evidence linking vagal activation to modulation of nociceptive processing[27]. Furthermore, vagal stimulation has been shown to promote gastrointestinal motility and accelerate the return of bowel function, which are key determinants of early recovery[28]. Improved autonomic stability and reduced symptom burden may in turn translate into higher quality of recovery, as measured by the QoR-15 scale[29]. Collectively, these secondary outcomes will provide complementary insights into the broader impact of taVNS on postoperative recovery and may support its integration into multimodal Enhanced Recovery After Surgery (ERAS) pathways.

Nevertheless, several limitations should be acknowledged. As a single-center study focusing on gynecologic laparoscopic procedures, the generalizability of the results to other surgical populations remains to be determined. In addition, this trial emphasizes clinical outcomes and does not include physiological measures of autonomic modulation such as heart rate variability or inflammatory biomarkers, which could provide mechanistic insights into the antiemetic effects of taVNS. Further multicenter studies incorporating objective autonomic or neurophysiological endpoints will be needed to validate and extend our findings.

In conclusion, this randomized, sham-controlled trial will evaluate the clinical effects and safety of perioperative taVNS as an adjunct to standard PONV prophylaxis in women undergoing gynecologic laparoscopic surgery. The findings may inform the design of larger studies and the potential role of auricular neuromodulation in multimodal perioperative care.

## Data Availability

No datasets were generated or analysed during the current study. All relevant data from this study will be made available upon study completion.

## Author contributions

Conceptualization and methodology: [RW, JJY, and LH]. Investigation and project administration: [RW, HY, HJL, JLD, YRY, YWZ]. Writing—original draft: [RW]. Writing—review and editing: [JJY and LH]. Supervision: [JJY and LH]. Funding acquisition: [LH]. All authors reviewed and approved the manuscript and agree to be accountable for their contributions and the integrity of the work.

## Acknowledgements

The authors thank the anesthesiology, nursing, and research teams for their support in developing and implementing the study.

## Data availability

This manuscript describes a trial protocol and does not report participant-level results. Following publication of the main trial findings, de-identified participant data, the data dictionary, relevant CRFs, and statistical analysis syntax may be made available upon reasonable request to the corresponding author, subject to participant consent, institutional approval, applicable data protection requirements, and an appropriate data-sharing agreement.

## Consent for publication

Not applicable. This protocol does not contain identifiable participant information.

## Dissemination

The findings will be submitted to a peer-reviewed journal and presented at scientific meetings. Participants will be offered a plain-language summary upon request. Results will also be communicated to relevant clinical departments to support evidence-informed discussion of perioperative symptom management.

